# Long-Acting Injectable Buprenorphine Use and Treatment Attribute Priorities Among U.S. Buprenorphine Prescribers: A National Survey

**DOI:** 10.64898/2026.02.01.26345319

**Authors:** Nicholas L. Bormann, Stephan Arndt, Tyler S. Oesterle

**Author notes:** <u>Corresponding author</u>: Nicholas L. Bormann, 200 1st St SW, Rochester, MN 55905. <u>Declarations of interest</u>: Oesterle provides consultative services to Koa Health and Frenelle Pharma. Oesterle has not received any compensation for these services. All other authors are employed by their respective institutions as noted. No other author discloses any financial and personal relationships with other people or organizations that could inappropriately influence or bias their work.

## Abstract

**Background:** Long-acting injectable buprenorphine (LAI-BUP) is safe and effective, however is dramatically underutilized in comparison to oral formulations. Little is known regarding how buprenorphine prescribers view LAI-BUP, and which medication attributes they prioritize when selecting treatment for opioid use disorder (OUD).

**Methods:** A secondary analysis of a national, cross-sectional online survey of U.S. physicians who prescribe buprenorphine for OUD was conducted. Respondents reported OUD caseload, LAI-BUP use, and the importance of medication attributes relevant to treatment selection (e.g., efficacy, safety, ease of administration, ease of prescribing, and administrative requirements). Providers were categorized as no LAI-BUP use or, among LAI-BUP prescribers, Low vs High use based on a median split. Group comparisons used chi-square (or Fisher’s exact) tests for categorical variables and Jonckheere–Terpstra tests for ordinal responses.

**Results:** Among 125 respondents, 39 (31.2%) reported no patients receiving LAI-BUP. The remaining 86 (68.8%) were LAI-BUP prescribers, split evenly into Low and High (n’s=43; 34.4%) groups using a median cut of 23.2%. LAI-BUP use did not differ meaningfully by specialty, region, or practice setting. Greater LAI-BUP use was reported by providers with larger OUD panels. Ratings of key medication attributes were uniformly high.

**Conclusions:** LAI-BUP remains underused, with uptake highest among clinicians managing larger OUD caseloads. Measured attitudes toward medication attributes did not explain these differences. Future work should assess clinic workflow, staffing, reimbursement, and REMS burden, testing targeted implementation strategies using mixed-methods trials. Identifying what shifts clinicians from no use to low and high use may guide scalable implementation interventions.

## Introduction

Across medicine, long-acting injectables (LAIs) have improved adherence and outcomes by eliminating daily dosing. In bipolar disorder, LAIs lower the risk of rehospitalization compared with oral therapy.^1^ In diabetes, weekly glucagon-like peptide-1 receptor agonists offer greater glycemic control and weight loss compared to daily oral formulations.^2^ In human immunodeficiency virus treatment, LAI antiretrovirals maintain viral suppression while improving adherence and patient satisfaction.^3^ These conditions regularly co-occur with opioid use disorder (OUD), where challenges of relapse and poor adherence are pronounced.

By reducing dosing frequency to once weekly, monthly, or longer, LAIs minimize the number of daily decisions required to continue medication. This simplification can enhance adherence and may reduce the cognitive burden associated with managing recovery. LAIs may also mitigate stigma associated with addiction pharmacotherapy, such as negative perceptions of methadone clinics or feelings of shame when filling prescriptions at pharmacies.^4^ Additionally, LAIs virtually eliminate the risk of lost, stolen, or diverted medication. These factors support their logical use in OUD treatment.

The Food and Drug Administration (FDA) has approved three LAIs for OUD: one opioid antagonist (extended-release naltrexone; XR-NTX) and two formulations of the partial agonist buprenorphine (extended-release buprenorphine; LAI-BUP). XR-NTX achieves relapse-free survival comparable to daily sublingual buprenorphine once initiated, but induction challenges limit its broader effectiveness.^5^ In contrast, buprenorphine can be started with only partial detoxification, and micro-induction protocols even allow for co-use of buprenorphine with the full agonist opioid, lowering induction barriers even further.^6^ Once on buprenorphine, it can then be transitioned to LAI-BUP.

Pivotal trials have established the efficacy and safety of LAI-BUP. Weekly and monthly LAI-BUP regimens yield substantially higher rates of opioid abstinence than placebo and are at least noninferior to daily sublingual buprenorphine for opioid-negative urine tests.^7-9^ Long-term safety studies report high treatment retention, significant rates of abstinence, and primarily injection-site adverse events.^7^ A recent systematic review of 18 studies concluded that LAI-BUP offers distinct advantages over oral formulations, including improved adherence and a reduced risk of diversion.^10^ Despite this evidence base, real-world use of LAI-BUP in the United States remains limited.

In a statewide analysis of Kentucky’s prescription drug monitoring data, between April 2019 and December 2020, only 879 of 71,114 individuals (1.2%) receiving buprenorphine were prescribed LAI-BUP. Kentucky subsequently removed the prior authorization requirement for Medicaid and expanded Risk Evaluation and Mitigation Strategy (REMS) certified sites, which approximately doubled the rate of LAI-BUP initiations.^11^ Within the Rhode Island Department of Corrections, just 2.5% of patients receiving buprenorphine were treated with LAI-BUP, even though its use was found to be feasible, well-tolerated, and showed no evidence of diversion.^12^ In contrast, Australia experienced rapid adoption of LAI-BUP following reimbursement reform, which provided universal coverage and streamlined prescribing and dispensing. LAI-BUP prescriptions there rose from 4.0% to 49.6% of all buprenorphine treatment from September 2019 to December 2022.^13^

Qualitative work highlights the complexity of patient and provider perspectives. Patients describe LAI-BUP benefits of increased freedom, less stigma, and fewer daily disruptions than daily or multiple-times-per-day dosing; however, they also experience injection discomfort, early withdrawal sensations, and reduced contact with clinic staff.^14-18^ Providers similarly report both enthusiasm and hesitation. They note potential advantages for individuals struggling with adherence and stability, but also cite limited experience, uncertainty around patient selection, medication costs, and administrative complexity as barriers to routine use.^19,20^ A key contributor to this complexity is the requirement that currently approved LAI-BUP products be dispensed and administered within a REMS program.^21^ The REMS exists because accidental or intentional intravenous administration of existing formulations can produce vascular occlusion, local tissue damage, or thromboembolic events due to product solidification upon contact with blood.^22,23^ These regulatory requirements introduce logistical challenges that can limit widespread availability.

Despite evidence supporting LAI-BUP, relatively little is known about how U.S. prescribers use it in practice or how they view potential new formulations designed to lessen current barriers. We conducted a secondary analysis of a national sample of buprenorphine prescribers to (1) describe patterns of LAI-BUP use across provider groups and (2) evaluate attitudes toward key medication attributes relevant to treatment selection.

## Materials and Methods

### Participants

This manuscript reports a secondary analysis of an existing, de-identified survey dataset. Respondents were recruited from the Sermo medical market research panel of verified healthcare professionals. Panel members categorized by Sermo as adult primary care (internal medicine, general practice, or family medicine), adult psychiatry, or addiction medicine were invited via an online screening survey to determine eligibility for the full research survey.

Eligible physicians were U.S. board-certified and reported providing direct care for ≥20 adults with OUD receiving medication treatment within the prior 6 months, personally initiating medications for OUD, and having 3–35 years of practice since completing residency. Recruitment quotas targeted 75 respondents from adult psychiatry and/or addiction medicine and 50 respondents from adult primary care.

Recruitment incorporated vendor-defined strata based on the proportion of buprenorphine prescriptions administered as a LAI-BUP, with specialty-specific thresholds. For adult primary care, the “higher LAI-BUP” stratum was defined as >5% LAI-BUP prescriptions and the “non/low LAI-BUP” stratum as ≤5%. For adult psychiatry/addiction medicine, the “higher LAI-BUP” stratum was defined as >10% LAI-BUP prescriptions and the “non/low LAI-BUP” stratum as ≤10%. Enrollment caps included a maximum of n=30 respondents in both the non/low LAI-BUP adult psychiatry/addiction medicine and the non/low LAI-BUP adult primary care subgroups.

Participants were incentivized to complete the screening and research surveys using standard market research rates based on estimated survey duration. Incentives were issued only after completion of the full research survey and validation of responses per panel quality-control procedures. Institutional Review Board for Human Subjects Research of Frenelle Pharma gave ethical approval for this work, and informed consent was obtained from all participants.

### Survey Instrument

The online survey included items assessing provider characteristics, OUD clinical caseload, use of LAI-BUP, and attitudes toward medication attributes relevant to treatment selection for OUD (e.g., perceived efficacy, safety, ease of administration, and ease of prescribing). Respondents in the non/low LAI-BUP pathway took approximately 20 minutes to complete the survey, while respondents in the higher LAI-BUP subgroup took approximately 30 minutes due to a greater number of questions asked.

### Statistical Analysis

Recruitment quotas used vendor-defined LAI-BUP prescribing strata. For this secondary analysis, respondents were reclassified using the proportion of each provider’s OUD panel receiving LAI-BUP to define three ordered groups. For each respondent, the percentage of OUD patients currently receiving an LAI-BUP was calculated. Providers with no LAI-BUP use were classified as the Zero group. Among providers with at least one LAI-BUP patient, a median split of this percentage was used to define the Low and High LAI-BUP use groups.

Many response variables were highly skewed. Therefore, chi-square tests (or Fisher’s exact tests when expected cell counts were small) were used for basic demographic categories, and Jonckheere-Terpstra tests were applied for ordinal responses. The Jonckheere-Terpstra method recognizes the ordinal nature of the provider groups (Zero, Low, or High) and tests for ordered responses.

## Results

A total of 1,100 providers were screened to obtain 125 participants, with no missing data. Reasons for exclusion were incomplete screening, failure to meet eligibility criteria, non-completion of the full survey, removal following quality-control checks, or quota closure. Of these, 39 (31.2%) reported no patients on LAI-BUP (Zero group). The remaining 86 patients (68.8%) were LAI-BUP prescribers, and, based on a median split at 23.2% of OUD patients receiving LAI-BUP, the High and Low groups were evenly divided into two groups of 43 patients (34.4%) each. Basic demographic and practice characteristics are shown in Table 1. The LAI-BUP usage groups did not differ meaningfully by specialty, region, or practice setting. However, the number of OUD patients cared for varied markedly, with those in the Zero group having the least, those in the Low group seeing over twice as many, and the High group seeing the most.

**Table 1.** Basic demographics.

| Variable | Zero<br>N = 39<sup>1</sup> | Low<br>N = 43<sup>1</sup> | High<br>N = 43<sup>1</sup> | p-value<sup>2</sup> |
| --- | --- | --- | --- | --- |
| **Primary specialty** |  |  |  | 0.24 |
| Adult Internal Medicine/General Practice/Family Medicine | 28 (72%) | 24 (56%) | 24 (56%) |  |
| Adult Psychiatry | 11 (28%) | 19 (44%) | 19 (44%) |  |
| **Region** |  |  |  | 0.39 |
| Midwest | 9 (23%) | 7 (16%) | 8 (19%) |  |
| Northeast | 11 (28%) | 13 (30%) | 15 (35%) |  |
| South | 5 (13%) | 1 (2.3%) | 7 (16%) |  |
| Southeast | 7 (18%) | 9 (21%) | 5 (12%) |  |
| West | 7 (18%) | 13 (30%) | 8 (19%) |  |
| **Geographic setting** |  |  |  | 0.40 |
| Rural | 5 (13%) | 1 (2.3%) | 2 (4.7%) |  |
| Suburban | 17 (44%) | 18 (42%) | 18 (42%) |  |
| Urban | 17 (44%) | 24 (56%) | 23 (53%) |  |
| **Years of practice** | 15.72 (9.17) | 10.47 (8.00) | 12.95 (8.72) | 0.21 |
| **Number of OUD patients on MAT** | 71.28 (67.17) | 167.07 (153.70) | 202.09 (206.08) | <0.001 |
| **Percent patients receiving LAI** | 0.00 (0.00) | 12.42 (6.49) | 36.10 (9.64) | <0.001 |

<sup>1</sup> n (%); Mean (SD)<sup>2</sup> Pearson's Chi-squared test; Fisher's exact test; Jonckheere-Terpstra test

Providers rated the importance of product attributes when selecting a medication for OUD. Table 2 summarizes these ratings for efficacy, safety, ease of administration, and ease of prescribing. Ratings were uniformly high, with no consistent differences across LAI-BUP groups.

**Table 2.** Efficacy, safety, ease of administration, and ease of prescribing.

| Variable | Zero<br>N = 39<sup>1</sup> | Low<br>N = 43<sup>1</sup> | High<br>N = 43<sup>1</sup> | p-value<sup>2</sup> |
| --- | --- | --- | --- | --- |
| Efficacy-related |  |  |  |  |
| Prevent relapse | 6.18 (1.05) | 6.12 (0.98) | 5.77 (1.17) | 0.092 |
| Reduce cravings | 6.23 (0.87) | 5.77 (1.15) | 5.72 (1.14) | 0.046 |
| Prevent overdose | 6.33 (0.96) | 6.28 (0.91) | 5.81 (1.33) | 0.10 |
| Block effects of illicit opioids | 5.62 (1.25) | 5.19 (1.22) | 5.37 (1.02) | 0.25 |
| Treat depression/ mood | 5.28 (1.17) | 4.63 (1.22) | 4.91 (1.27) | 0.23 |
| Safety-related |  |  |  |  |
| No boxed warning or REMS | 5.13 (1.08) | 4.93 (1.33) | 4.86 (1.25) | 0.42 |
| Low abuse/diversion potential | 5.77 (1.20) | 5.70 (1.12) | 5.77 (1.13) | 0.93 |
| Tolerability | 5.59 (1.02) | 5.44 (1.24) | 5.56 (1.14) | 0.90 |
| Low interactions w/ other meds | 5.67 (1.08) | 5.23 (1.13) | 5.49 (1.05) | 0.51 |
| Ability to be reversed | 5.31 (1.30) | 4.98 (1.41) | 5.07 (1.12) | 0.56 |
| Ease of administration |  |  |  |  |
| Patient preference | 5.41 (1.29) | 5.28 (1.24) | 5.47 (1.12) | 0.86 |
| Formulation | 5.00 (1.26) | 4.74 (1.16) | 5.21 (1.01) | 0.47 |
| Ability to be weaned off | 5.38 (1.25) | 5.07 (1.45) | 5.23 (1.25) | 0.65 |
| Retention on treatment | 6.03 (1.16) | 5.91 (0.84) | 5.58 (1.31) | 0.080 |
| Access at pharmacy | 5.31 (1.34) | 5.33 (1.17) | 5.44 (1.26) | 0.61 |
| Ease of prescribing |  |  |  |  |
| Ease of induction | 5.49 (1.30) | 5.35 (1.13) | 5.47 (1.10) | 0.89 |
| Insurance coverage | 5.54 (1.31) | 5.16 (1.48) | 5.47 (1.22) | 0.78 |
| No special training to prescribe | 4.64 (1.66) | 5.07 (1.24) | 5.02 (1.50) | 0.24 |

<sup>1</sup> Mean (SD)
<sup>2</sup> Jonckheere-Terpstra test

## Discussion

This survey of buprenorphine prescribers found that LAI-BUP remains underused, with nearly one-third of respondents reporting no use. A unique finding was that providers with larger OUD caseloads were more likely to prescribe LAI-BUP. In contrast, LAI-BUP use did not differ meaningfully by specialty, region, or practice setting. Priorities around key medication attributes were also similar across groups. Together, these findings suggest that factors beyond those measured in this survey may be influencing LAI-BUP utilization.

These practice patterns contrast with the robust evidence base supporting LAI-BUP. In a phase 3 randomized controlled trial, two monthly dosing levels of LAI-BUP produced markedly higher rates of opioid abstinence, averaging 41.3-42.7% of weeks opioid-negative compared with 5.0% in the placebo group. Reported adverse effects were primarily injection-site reactions, with no increase in serious adverse events.^8^ In a 48-week international safety study, weekly and monthly formulations demonstrated high retention (73.6%) and substantial abstinence, with opioid-negative urine tests in 63.0% of new patients and 82.8% of those switched from sublingual buprenorphine, along with high patient tolerability and satisfaction.^7^ In a 24-week double-blind comparison trial, LAI-BUP was noninferior to daily sublingual buprenorphine for sustained abstinence and statistically superior when considering the overall distribution of opioid-negative urine samples.^9^ Against this backdrop, the modest rates of LAI-BUP use in this sample highlight a gap between evidence and implementation.

Provider perspectives have been less characterized than patients. In a Swedish qualitative study of 10 physicians who prescribe opioid agonist treatment and an additional 41 health care staff involved (i.e., nurses, counselors, clinical caretakers), they overall felt that LAI-BUP had significant advantages. However, their views evolved from initially being appropriate mainly for more stable patients to actually being particularly helpful for individuals struggling with adherence and recovery.^19^ In a U.S. study of 20 members of multidisciplinary addiction teams, including physicians, nurse practitioners, social workers, and peer recovery specialists, barriers to LAI-BUP use included limited experience, uncertainty around patient selection, medication cost, and administrative complexity.^20^ The REMS is a key driver of this complexity. The high endorsement of a REMS-free product in this survey aligns with these qualitative findings and highlights how regulatory and procedural features may limit uptake even when clinical attitudes are favorable.

Despite assessing perceptions of efficacy, safety, administrative burden, and ease of prescribing, this study did not identify specific attitudinal factors that clearly distinguished zero, low, and high LAI-BUP prescribers or functioned as discrete “levers” to move clinicians along that continuum. This may reflect the level at which questions were asked. Future surveys may assess additional operational domains, such as the impact of injection clinic staffing, availability of clinical space and medication storage, reimbursement and billing workflows, and perceptions of regulatory risk associated with REMS participation on rates of LAI-BUP use. Mixed-methods work that links prescribing data to interviews or focus groups could help identify specific changes, such as changes to REMS, pharmacy-based administration, or integration into existing injection clinics, that may increase the likelihood of a provider shifting from non-use to occasional prescribing and from occasional to routine use.

The association between higher OUD caseload and greater LAI-BUP use may also have policy relevance. Historically, the X-waiver system placed caps on the number of patients a clinician could treat with buprenorphine, potentially limiting the development of high-volume OUD practices with the infrastructure needed to support LAI-BUP.^24^ The removal of those limits may allow some prescribers to build larger OUD panels, gain experience with injectable formulations, and justify the effort required to implement LAI-BUP workflows.^24^ Future research should investigate whether increasing buprenorphine caseloads over time are associated with subsequent growth in LAI-BUP use, and whether targeted implementation strategies in high-volume settings can accelerate this trajectory.

This study does have limitations. Rates of LAI-BUP use were self-reported, and clinicians may have over- or under-estimated panel size. The cross-sectional design limits the ability to gain a deeper understanding of the relationships between OUD caseload, experience, and LAI-BUP use. However, this was the largest sample of buprenorphine prescribers on LAI-BUP. Although the sample was recruited from across the U.S., this may not be generalizable to all buprenorphine prescribers or those in other countries.

## Conclusions

While LAI-BUP has strong efficacy and safety data, it remains underutilized among surveyed buprenorphine prescribers. A key finding was that uptake was closely tied to OUD caseload rather than to negative views of injectable treatment, which may reflect increasing comfort with the intervention or the provider adapting to an intervention patients are interested in. This pattern suggests that administrative burden or workflow constraints, rather than negative views of injectable treatment, may be contributors of low uptake. While logistical and regulatory features may be key constraints, these are modifiable, and countries could follow policy successes like that seen in Australia. Efforts to expand LAI-BUP use will likely require increased education, advocacy, and the development of formulations and policies that simplify delivery while preserving safety and effectiveness.

## Data Statement

All data discussed in this manuscript are available upon reasonable request to the authors.

## Role of funding source

The study was funded by Frenelle Pharma, including developing the survey and funding participant remuneration. The sponsor did not compensate the authors and had no role in manuscript writing. The authors performed the analyses and interpreted the findings independently.

## Data Availability

All data discussed in this manuscript are available upon reasonable request to the authors.

## Acknowledgments

We thank Frenelle PBC for funding the study, collecting the raw data, and providing the authors access to this data.

## Notes

### Author Declarations

Institutional Review Board for Human Subjects Research of Frenelle Pharma gave ethical approval for this work, and informed consent was obtained from all participants.

